# Application of Machine Learning and Deep Learning in diagnosis Covid19

**DOI:** 10.64898/2026.11.01.26367023

**Authors:** Loubna Merchi, Cheima Boucheham, Abdesslem Layeb, Asma Derouiche

**Affiliations:** Department of Fundamental Computing and its Applications (IFA), Faculty of New Technologies of Information and Communication (NTIC), University of Abdelhamid Mehri – Constantine 2, Constantine, Algeria; Internal Medicine Department, University Hospital Centre (UCH) of Constantine, Algeria

**Keywords:** COVID-19, prognosis, machine learning, deep learning, feature selection, multilayer perceptron, clinical decision support

## Abstract

Predicting the progression of COVID-19 in hospitalised patients was an important challenge during the pandemic, particularly when healthcare resources were limited. This study investigates the use of machine learning (ML) and deep learning (DL) models to predict three clinical outcomes: death, admission to the intensive care unit (ICU), and clinical status (improvement or worsening).

The study uses two patient datasets: an Algerian cohort of 80 patients from the Internal Medicine Department of the University Hospital Centre of Constantine, collected between July and August 2020, and a French cohort of 99 patients collected between March and May 2020. Five ML classifiers were evaluated: logistic regression (LR), k-nearest neighbours (KNN), classification and regression trees (CART), Naive Bayes (NB), and support vector machines (SVM). Their performance was examined in combination with three feature-selection methods: analysis of variance (ANOVA), mutual information (MI), and recursive feature elimination (RFE). The results were also compared with multilayer perceptron (MLP) models using one to five hidden layers. Model evaluation included 30 repetitions of five-fold cross-validation on the training data, followed by evaluation on a separate hold-out test set.

The results showed that selecting 20 features generally provided better cross-validated accuracy than selecting 30 features or using the full feature set. The best-performing classifier–feature-selection combination varied across the six dataset–outcome tasks. LR combined with RFE was selected for three tasks, LR combined with ANOVA for one task, and CART combined with ANOVA for two tasks. The selected ML models achieved test-set accuracies ranging from 0.47 to 0.95 and higher accuracy than the average MLP results in five of the six tasks.

These findings suggest that ML methods can be useful for exploring clinical outcomes in hospitalised COVID-19 patients. However, the small sample sizes and limitations of the available data mean that the results should be interpreted with caution. Further evaluation using larger and independent patient datasets is needed before these models can be considered for clinical use.

## 1 Introduction

### Research context

Over the years, the world has experienced several pandemics that have caused significant human and economic losses. However, the COVID-19 pandemic presented a major challenge to healthcare systems worldwide, particularly in the diagnosis and management of patients. Despite advances in medicine and healthcare, hospitals struggled to manage the large number of patients, resulting in significant human losses.

In this context, Data Science and Machine Learning offer opportunities to support healthcare professionals in assessing patients’ conditions and predicting the possible progression of diseases. These technologies can help improve healthcare processes by collecting, analysing and interpreting patient data. More generally, the growing use of Data Science, Artificial Intelligence and Machine Learning in healthcare has contributed to improving diagnostic and treatment processes, supporting timely medical decisions and increasing the efficiency of healthcare services.

### Problem statement and motivation

Traditional healthcare resource management, overloaded medical services and the lack of appropriate computer systems and medical technologies can affect the quality of care provided by healthcare institutions. In addition, healthcare professionals may lack the tools and data-science expertise needed to visualise and interpret medical data and make use of information from patients’ medical histories.

During the COVID-19 pandemic, the absence of sufficient prognostic models made it difficult to anticipate which patients were likely to recover and be discharged and which might deteriorate or die and require more specialised treatment. This situation highlighted the need for predictive approaches that could help healthcare professionals assess patients’ conditions and better manage available resources.

### Objectives and contributions

The objective of this work is to develop a predictive approach for assessing the final clinical state of hospitalised COVID-19 patients using Machine Learning and Deep Learning techniques. Feature-selection methods are also employed to identify the patient features that are most relevant to the assessment of their conditions.

The study investigates five Machine Learning classifiers: Logistic Regression (LR), K-Nearest Neighbours (KNN), Classification and Regression Tree (CART), Naïve Bayes (NB) and Support Vector Machine (SVM). It also examines an Artificial Neural Network (ANN) as a Deep Learning model. Three feature-selection techniques are considered: Recursive Feature Elimination (RFE), Analysis of Variance (ANOVA) and Mutual Information (MI).

The experimental study is conducted using two datasets. The first is an Algerian dataset developed in cooperation with the Internal Medicine Department of the University Hospital Centre (UCH) of Constantine. The second is a French dataset obtained from previous research by Billoir et al. (2021). Through this comparative study, we aim to evaluate the different approaches and identify the patient features that may be important for assessing clinical conditions.

### Paper organisation

Section 2 reviews related work; Section 3 describes data and methods; Section 4 reports results; Section 5 discusses findings and limitations; Section 6 concludes.

## 2 Related Work

Machine learning (ML) and deep learning (DL) have been increasingly applied to healthcare problems, including disease diagnosis and the prediction of clinical outcomes. Their performance can vary depending on the dataset, the selected features and the algorithms used. Previous studies have explored these approaches in different medical fields, including diabetes prediction, heart disease classification and COVID-19 analysis. Table 1 summarises the studies discussed in this work, with performance figures as reported by the original authors.

**Table 1:**
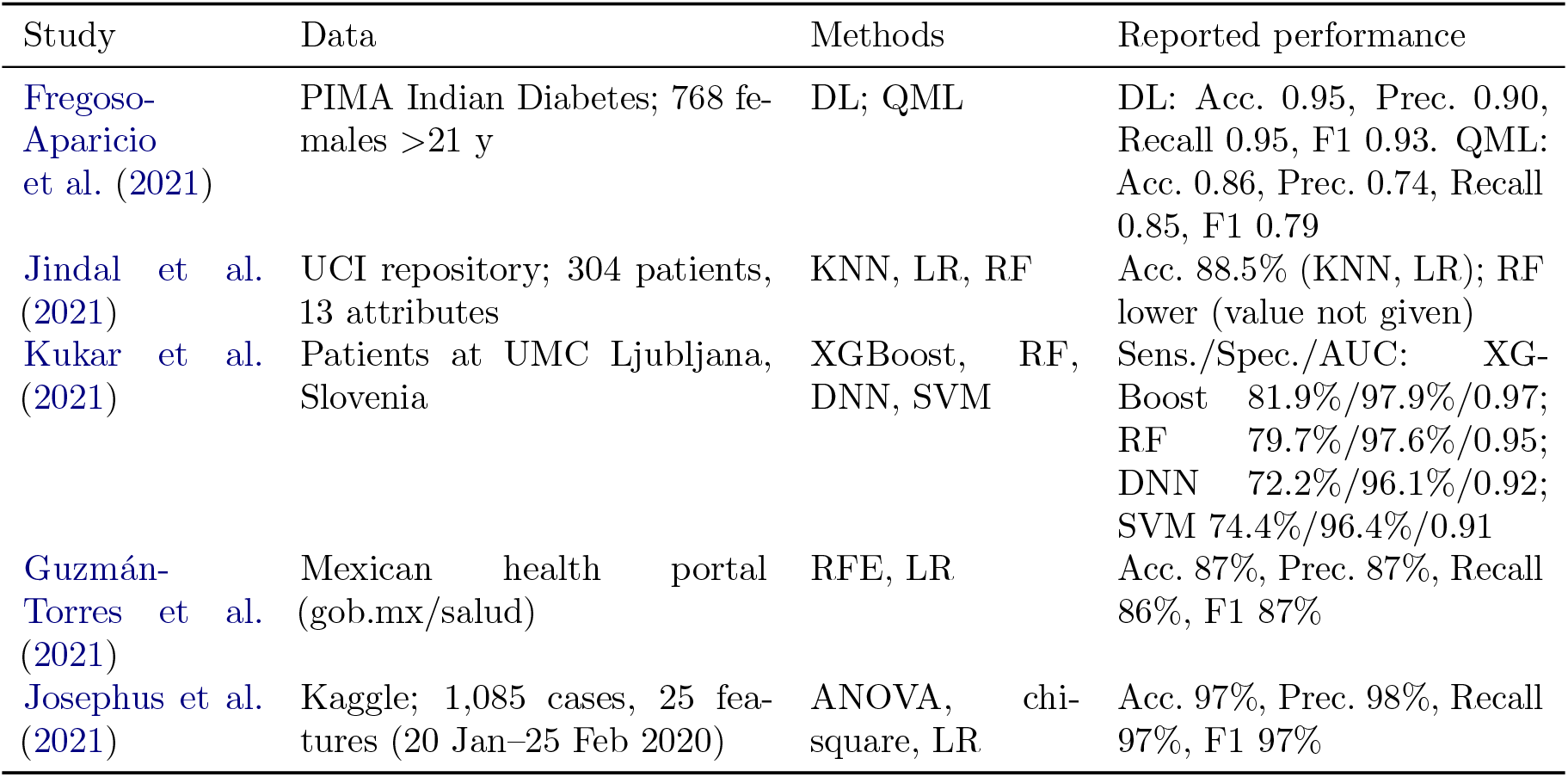
Related works.

| Study | Data | Methods | Reported performance |
|---|---|---|---|
| Fregoso-Aparicio et al. (2021) | PIMA Indian Diabetes; 768 females >21 y | DL; QML | DL: Acc. 0.95, Prec. 0.90, Recall 0.95, F1 0.93. QML: Acc. 0.86, Prec. 0.74, Recall 0.85, F1 0.79 |
| Jindal et al. (2021) | UCI repository; 304 patients, 13 attributes | KNN, LR, RF | Acc. 88.5% (KNN, LR); RF lower (value not given) |
| Kukar et al. (2021) | Patients at UMC Ljubljana, Slovenia | XGBoost, RF, DNN, SVM | Sens./Spec./AUC: XGBoost 81.9%/97.9%/0.97; RF 79.7%/97.6%/0.95; DNN 72.2%/96.1%/0.92; SVM 74.4%/96.4%/0.91 |
| Guzmán-Torres et al. (2021) | Mexican health portal (gob.mx/salud) | RFE, LR | Acc. 87%, Prec. 87%, Recall 86%, F1 87% |
| Josephus et al. (2021) | Kaggle; 1,085 cases, 25 features (20 Jan–25 Feb 2020) | ANOVA, chi-square, LR | Acc. 97%, Prec. 98%, Recall 97%, F1 97% |

Fregoso-Aparicio et al. (2021) presented a systematic review of machine learning and deep learning models for type 2 diabetes prediction. Their work examined the application of different predictive approaches in this field. Jindal et al. (2021) investigated the use of machine learning algorithms to predict heart disease using a dataset of 304 patients with 13 medical attributes obtained from the UCI repository. They compared K-nearest neighbors (KNN), logistic regression (LR) and random forest (RF). KNN and LR achieved the highest reported accuracy, at 88.5%.

Several studies have also applied machine learning techniques to COVID-19-related problems. Kukar et al. (2021) investigated the use of routine laboratory and clinical indicators to distinguish COVID-19-positive patients from negative participants at the University Medical Centre Ljubljana in Slovenia. They evaluated XGBoost, random forest, deep neural networks (DNN) and support vector machines (SVM). XGBoost achieved the best reported performance, with a sensitivity of 81.9%, specificity of 97.9% and AUC of 0.97.

Guzmán-Torres et al. (2021) used supervised machine learning to identify conditions associated with an increased risk of death among COVID-19 patients in Mexico. Their approach combined recursive feature elimination (RFE) with logistic regression to select relevant variables and predict patient mortality. The study reported an accuracy of 87%.

Similarly, Josephus et al. (2021) investigated mortality prediction among COVID-19 patients using a dataset of 1,085 cases and 25 features. They applied ANOVA and chi-square feature-selection techniques with logistic regression. The reported accuracy was 97%.

### Research Gap and Study Motivation

The reviewed studies show that different machine learning and deep learning methods can achieve different results depending on the dataset and the prediction task. There is no single method that performs best in all cases. Moreover, the studies highlight the importance of data preprocessing and feature selection. Therefore, this work investigates several machine learning classifiers, feature-selection techniques, and deep learning models using two datasets from different countries, with the aim of identifying the most suitable approaches for predicting COVID-19 patient outcomes.

## 3 Materials and Methods

### 3.1 Overview

The objective of this study is to develop a predictive analytics approach for assessing the final clinical state of COVID-19 patients using Machine Learning (ML) and Deep Learning (DL) techniques. The proposed approach consists of three main stages: collecting patient data, preprocessing the data, and applying trained classification models to predict the patient’s condition.

Two types of models are investigated: ML classifiers combined with feature-selection methods and a DL model. The experiments are conducted separately on the Algerian and French datasets to identify the most suitable models for predicting each clinical outcome.

The overall workflow of the proposed approach is illustrated in Figure 1.

**Figure 1:**
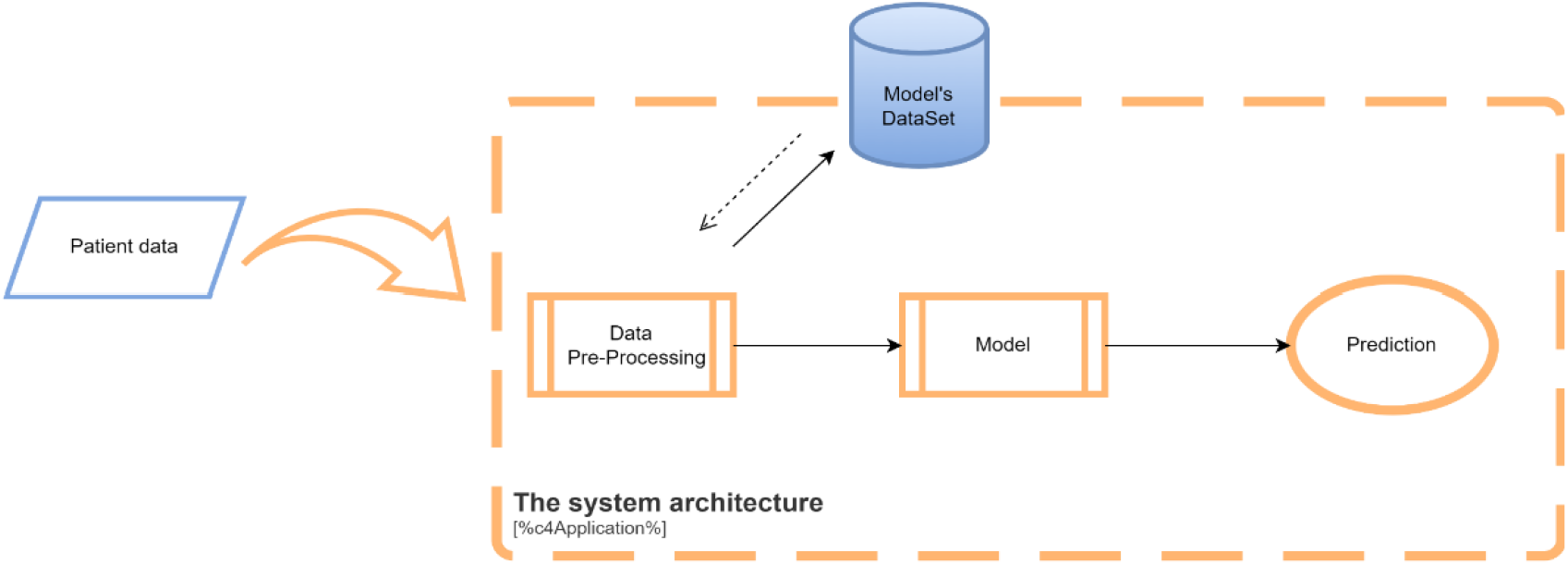
Predictive analytic system for Covid-19 patients

The ML and DL model-building workflows are shown in Figure 2 and 3.

**Figure 2:**
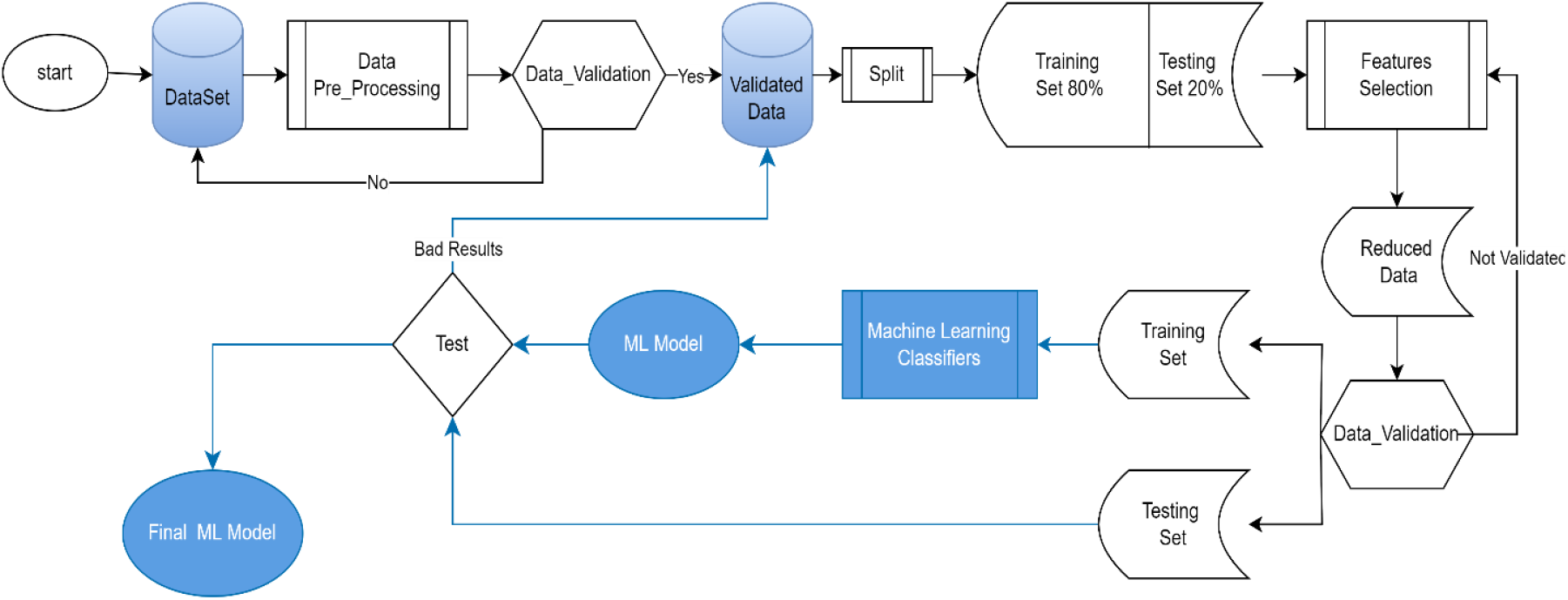
ML Model-building workflow.

**Figure 3:**
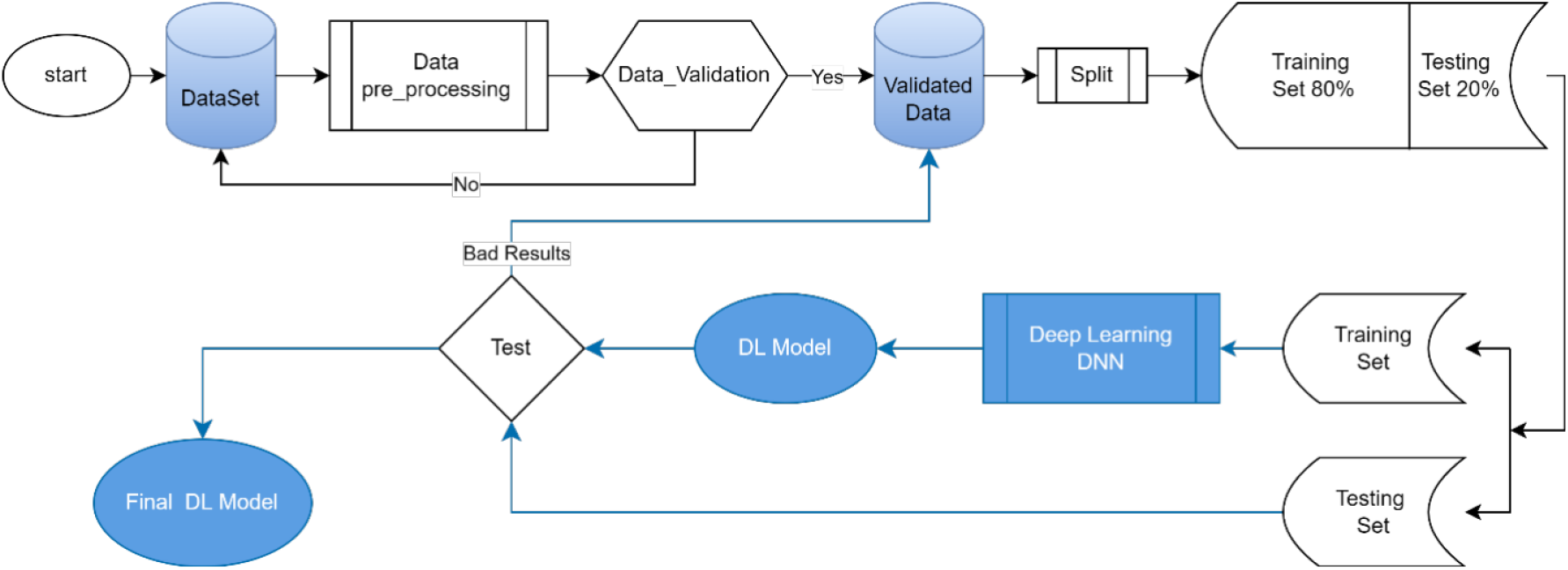
DL Model-building workflow.

### 3.2 Datasets

Two retrospective datasets were used (Table 2; full attribute listings in Appendix A).

**Table 2:**
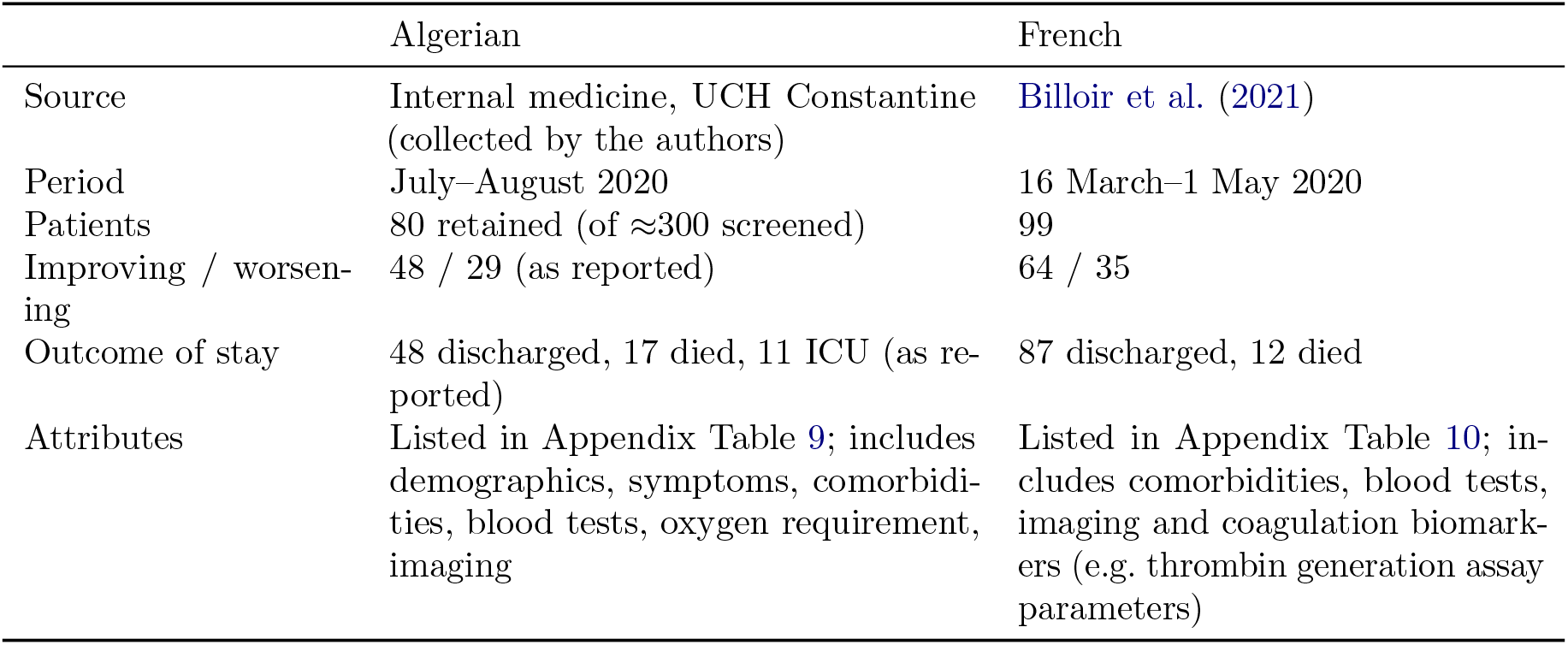
Characteristics of the Algerian and French datasets.

|  | Algerian | French |
|---|---|---|
| Source | Internal medicine, UCH Constantine (collected by the authors) | Billoir et al. (2021) |
| Period | July–August 2020 | 16 March–1 May 2020 |
| Patients | 80 retained (of ≈300 screened) | 99 |
| Improving / worsen-ing | 48 / 29 (as reported) | 64 / 35 |
| Outcome of stay | 48 discharged, 17 died, 11 ICU (as re-ported) | 87 discharged, 12 died |
| Attributes | Listed in Appendix Table 9; includes demographics, symptoms, comorbidi-ties, blood tests, oxygen requirement, imaging | Listed in Appendix Table 10; in-cludes comorbidities, blood tests, imaging and coagulation biomark-ers (e.g. thrombin generation assay parameters) |

#### Algerian dataset

The dataset was compiled by the authors from the confidential archive of the internal medicine department of UCH Constantine, with the permission of the head of department and under the supervision of Dr. Derouiche.A. About 300 medical records of COVID-19 patients were screened, and 80 were retained because the others lacked information. Records contain lung imaging findings, blood tests, symptoms and other clinical information for the period July–August 2020; values were recorded at the date closest to admission to the department. Patients were labelled clinically “worsening” if transferred to the ICU or if they died, and “improving” otherwise.

#### French dataset

This dataset was taken from the research conducted by (Billoir et al., 2021), which investigated coagulation biomarkers to assess clinical deterioration in SARS-CoV-2 infection. We chose to work with this dataset because it was available and contained almost complete values. It includes 99 patients with COVID-19 hospitalised in dedicated COVID-19 medical units between 16 March and 1 May 2020. The aim of our study was to investigate the association between coagulation and COVID-19 progression. Patients were considered clinically “worsening” if they were transferred to the intensive care unit (ICU) or died, and clinically “improving” otherwise. Of the 99 patients, 64 were clinically “improving” and 35 were registered as clinically “worsening”; 87 were discharged and the remaining 12 were declared dead.

#### Outcomes

For both datasets three binary targets were modelled separately: *Death* (1 = died), *ICUadmission* (1 = admitted to ICU) and *Clinical* (1 = improvement, 0 = worsening).

### 3.3 Data pre-processing

The same steps were applied to both datasets:

1. *Outliers:* abnormal values were removed using plausible bounds for each attribute defined with the help of a doctor.
2. *Missing data:* columns or rows with more than 50% missing values were removed; remaining missing values were imputed with scikit-learn’s KNNImputer.
3. *Encoding:* categorical variables were binarised (e.g. Yes = 1, No = 0) and ordinal variables encoded accordingly.
4. *Standardisation:* each feature *x* was rescaled as

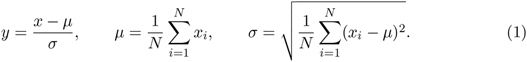

As shown in Figures 2 and 3, the preprocessing steps were performed before the training/test split.

### 3.4 Data split and feature sets

Each dataset was split into two sets: 80% for training and 20% for testing. The training set was used to train the models, while the testing set was used to evaluate their performance.

### 3.5 Machine learning models

Five machine learning classifiers were employed: logistic regression (LR; solver = liblinear, multi class = ovr), k-nearest neighbours (KNN; number of neighbours = 3), naïve Bayes (NB), support vector machine (SVM; gamma = auto), and classification and regression tree (CART).

Three feature-selection methods were employed: analysis of variance (ANOVA) and mutual information (MI), each evaluated with two different numbers of selected features (*N_fs_* = 20 and *N_fs_* = 30), and recursive feature elimination (RFE), using logistic regression as the estimator and a step size of 1.

The model-building process consisted of several experiments. First, the five classifiers (LR, KNN, CART, NB and SVM) were trained without feature selection to evaluate their performance using all available features. Next, ANOVA and MI were combined with the classifiers using 20 and 30 selected features. Finally, RFE was applied as a feature-selection method without fixing the number of selected features. This process resulted in 15 classifier/feature-selection combinations, as presented in Table 3.

**Table 3:** The 15 classifier/feature-selection pairs.

| ANOVA | MI | RFE |
|---|---|---|
| ANOVA, LR | MI, LR | RFE, LR |
| ANOVA, KNN | MI, KNN | RFE, KNN |
| ANOVA, CART | MI, CART | RFE, CART |
| ANOVA, NB | MI, NB | RFE, NB |
| ANOVA, SVM | MI, SVM | RFE, SVM |

#### Training the models

All models were trained 30 times using cross-validation on the training set. In this process, the input data were divided into (k) subsets. The model was trained on (k-1) subsets and evaluated on the remaining subset. This process was repeated (k) times, with a different subset reserved for evaluation each time. The performance of each training run was evaluated using accuracy, specificity, sensitivity, precision and F1-score.

#### Selecting the best model

The selection of the best model was based on the mean and standard deviation of the 30 training performance measurements, including accuracy, specificity, sensitivity, F1-score and precision. The model with the highest mean accuracy and the lowest standard deviation of accuracy was selected as the best model, while the other performance metrics were also taken into consideration.

#### Final evaluation

The selected model was evaluated on the testing set to assess its classification performance. The evaluation was based on accuracy, sensitivity, specificity, precision, F-score, the confusion matrix and the area under the ROC curve (AUC). The ROC curve illustrates the relationship between the true positive rate (TPR) and the false positive rate (FPR).

### 3.6 Deep learning model

A multilayer feed-forward perceptron (MLP) was used to build the deep learning model. The model was implemented using the Keras Sequential API, with ReLU as the activation function and the Adam optimiser to update the network weights during training. The MLP consists of input and output layers, with one or more hidden layers. Different numbers of hidden layers were evaluated to identify the configuration that achieved the best accuracy.

#### Evaluation and selection

The MLP was evaluated using different numbers of hidden layers. Its performance was assessed using loss, accuracy, F1-score, sensitivity, precision and specificity. The number of hidden layers that achieved the best accuracy was selected for the final model.

### 3.7 Evaluation metrics

The performance of the classification models was evaluated using accuracy, precision, sensitivity, specificity, and F1-score. These metrics were calculated based on the confusion matrix, where TP, TN, FP, and FN represent true positives, true negatives, false positives, and false negatives, respectively.

Accuracy measures the proportion of correctly classified observations:

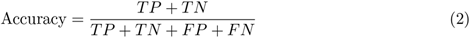

Precision measures the proportion of correctly predicted positive observations:

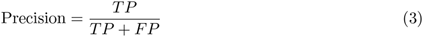

Sensitivity, also known as recall, measures the proportion of actual positive observations correctly identified:

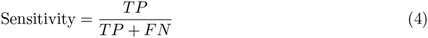

Specificity measures the proportion of actual negative observations correctly identified:

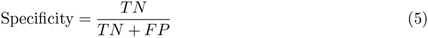

The F1-score combines precision and sensitivity through their harmonic mean:

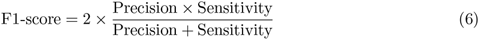

### 3.8 Statistical comparison and implementation

The Friedman test was used to evaluate and compare the results of the machine learning classifier/feature-selection combinations for each dataset and identify the best-performing combination.

The experiments were implemented in Python 3.9 using the Anaconda distribution and Spyder IDE. The libraries used included scikit-learn, Keras, TensorFlow, Matplotlib, Pandas, NumPy, Seaborn and openpyxl. The experiments were conducted on an HP ProBook laptop equipped with an Intel Core i7-6600U processor (2.60 GHz, 2.81 GHz), 8 GB of memory, a 500 GB HDD and Intel UHD Graphics 620, running Windows 10 64-bit.

## 4 Results

### 4.1 Cross-validation results

The cross-validation results showed that selecting 20 features generally gave better results than selecting 30 features or using all features. The best classifier and feature-selection combination varied depending on the dataset and prediction outcome. Table 4 presents the selected combinations and their corresponding mean cross-validated accuracy and standard deviation over 30 repetitions of five-fold cross-validation.

**Table 4:** Selected classifier and feature-selection pairs and their cross-validated accuracy (mean *±* SD).

| Dataset | Outcome | Selected pair | CV accuracy (mean ± SD) |
|---|---|---|---|
| Algerian | Death | CART + ANOVA | 1.00 ± 0.00 |
| Algerian | Clinical | LR + ANOVA | 0.79 ± 0.03 |
| Algerian | ICU admission | LR + RFE | 0.92 ± 0.03 |
| French | Death | LR + RFE | 0.94 ± 0.01 |
| French | Clinical | LR + RFE | 0.74 ± 0.03 |
| French | ICU admission | CART + ANOVA | 0.97 ± 0.02 |

### 4.2 Hold-out test performance of the selected ML models

Table 5 presents the hold-out test performance of the selected ML models, together with the confusion-matrix counts shown in Figure 4. The test sets were small, containing between 13 and 20 patients. Moreover, each confusion matrix included only two to six positive cases for Death and ICU admission. Therefore, the reported metrics should be interpreted with caution, as the classification of a single patient can substantially affect the results.

**Figure 4:**
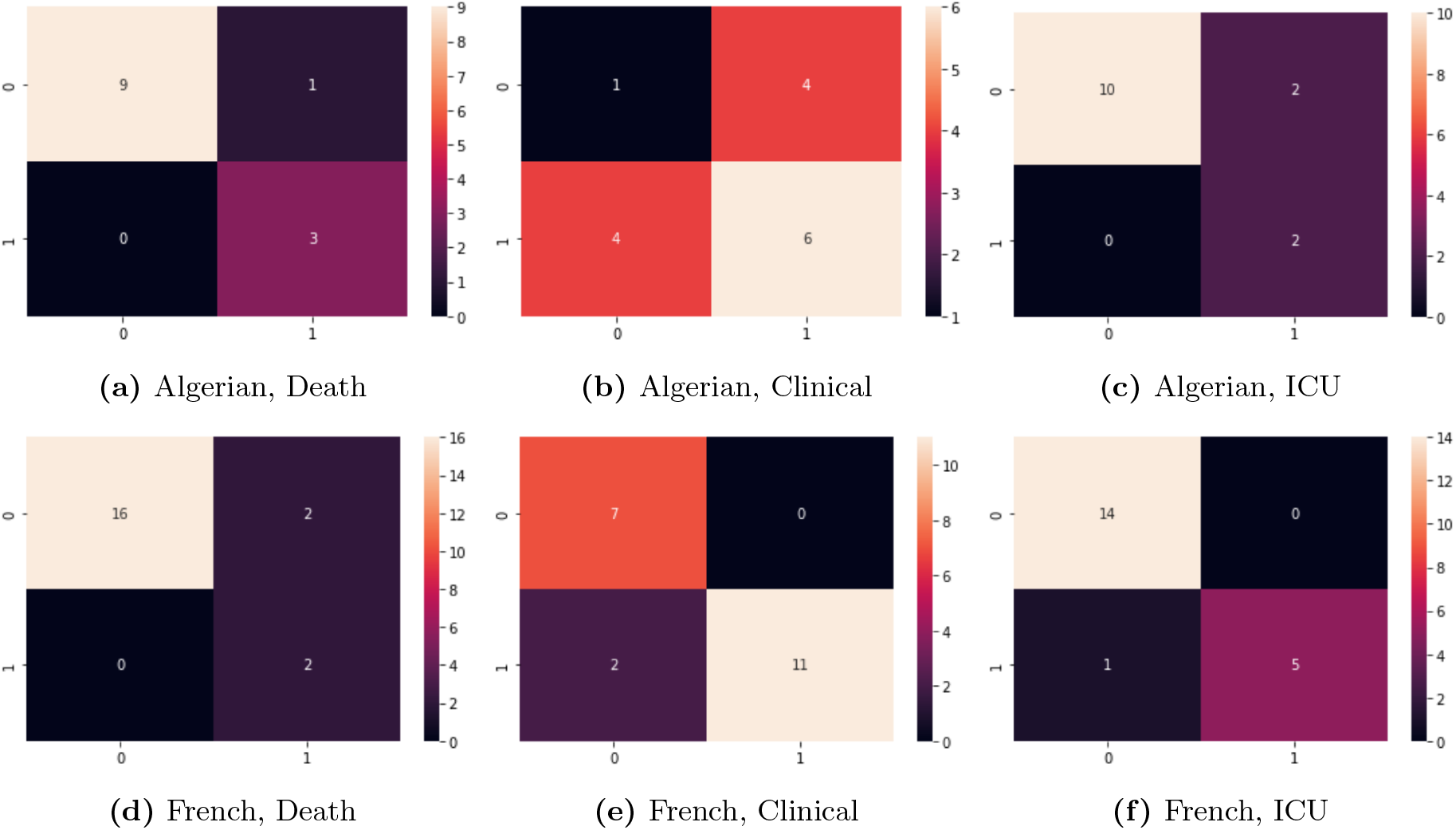
Confusion matrices of the selected ML models on the test sets

**Table 5:** Hold-out test performance of the selected ML models.

| Dataset | Outcome | Selected model | *n* | TN | FP | FN | TP | Accuracy | Sensitivity | Specificity | Precision | F1 | AUC |
|---|---|---|---|---|---|---|---|---|---|---|---|---|---|
| Algerian | Death | CART + ANOVA | 13 | 9 | 1 | 0 | 3 | 0.92 | 1.00 | 0.90 | 0.75 | 0.86 | 0.95 |
| Algerian | Clinical | LR + ANOVA | 15 | 1 | 4 | 4 | 6 | 0.47 | 0.60 | 0.20 | 0.60 | 0.60 | 0.40 |
| Algerian | ICU admission | LR + RFE | 14 | 10 | 2 | 0 | 2 | 0.86 | 1.00 | 0.83 | 0.50 | 0.67 | 0.92 |
| French | Death | LR + RFE | 20 | 16 | 2 | 0 | 2 | 0.90 | 1.00 | 0.89 | 0.50 | 0.67 | 0.94 |
| French | Clinical | LR + RFE | 20 | 7 | 0 | 2 | 11 | 0.90 | 0.85 | 1.00 | 1.00 | 0.92 | 0.92 |
| French | ICU admission | CART + ANOVA | 20 | 14 | 0 | 1 | 5 | 0.95 | 0.83 | 1.00 | 1.00 | 0.91 | 0.92 |

The Algerian models achieved accuracies of 0.92 for Death, 0.47 for Clinical status, and 0.86 for ICU admission. For the French dataset, the corresponding accuracies were 0.90, 0.90, and 0.95, respectively. The Algerian Clinical model showed the lowest performance, with an accuracy of 0.47 and an AUC of 0.40.

The confusion matrices also provide further insight into the classification results. All six models correctly identified the positive cases for Death and Algerian ICU admission, resulting in a sensitivity of 1.00 for these outcomes. However, their specificity values varied, indicating differences in their ability to correctly identify negative cases. In particular, the Algerian Clinical model had a specificity of 0.20, showing difficulty in correctly classifying patients with the negative clinical outcome. By contrast, the French Clinical and ICU admission models achieved a specificity of 1.00.

The ROC curve analysis yielded AUC values of 0.95, 0.40, and 0.92 for the Algerian Death, Clinical, and ICU admission models, respectively, and 0.94, 0.92, and 0.92 for the corresponding French models ( Figure 7).

### 4.3 Deep learning results

The deep learning models were implemented using multilayer perceptrons (MLPs) with one to five hidden layers. Figure 5 presents the validation accuracy across epochs for one run of each configuration and prediction task. In these individual runs, the highest validation accuracies were 1.00, 0.67, and 0.93 for the Algerian Death, Clinical, and ICU admission models, respectively. For the French dataset, the corresponding values were 0.95, 0.80, and 0.90. These results were obtained with five, three, and three hidden layers for the Algerian models, and two, one, and three hidden layers for the French models, respectively.

**Figure 5:**
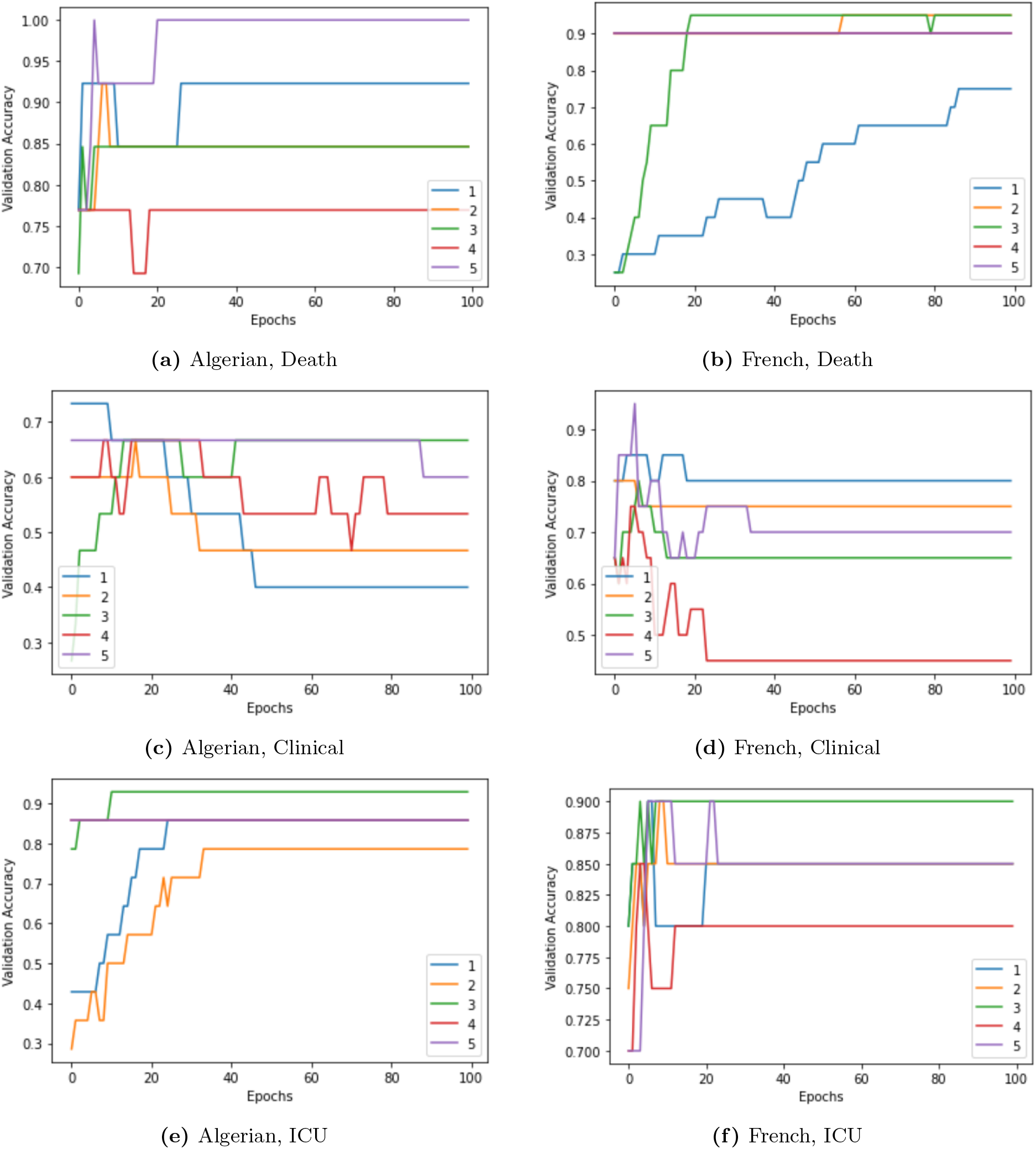
Validation accuracy across epochs for MLP models with different numbers of hidden layers across the three prediction outcomes in the Algerian and French datasets.

The selected single-run models generally showed higher performance during training than during validation, indicating possible overfitting. For example, the Algerian Clinical model achieved a training accuracy of 1.00, compared with a validation accuracy of 0.67. Since neural-network training can produce different results across runs, each hidden-layer configuration was subsequently evaluated over 30 runs. Table 6 summarizes the mean test accuracy and standard deviation for each configuration.

**Table 6:** Mean test accuracy (± standard deviation) of MLP models over 30 runs, according to the number of hidden layers, for the Algerian and French datasets.

| Dataset | Outcome | 1 HL | 2 HL | 3 HL | 4 HL | 5 HL |
|---|---|---|---|---|---|---|
| Algerian | Death | 0.78±0.08 | **0.81±0.08** | 0.79±0.11 | 0.75±0.12 | 0.72±0.10 |
| Algerian | Clinical | 0.39±0.09 | 0.43±0.11 | 0.49±0.11 | **0.51±0.11** | 0.49±0.17 |
| Algerian | ICUadmission | 0.75±0.10 | **0.81±0.07** | 0.78±0.13 | 0.78±0.11 | 0.80±0.14 |
| French | Death | 0.87±0.04 | **0.89±0.06** | **0.89±0.07** | 0.86±0.09 | 0.86±0.08 |
| French | Clinical | 0.75±0.08 | **0.76±0.08** | 0.72±0.10 | 0.68±0.11 | 0.65±0.09 |
| French | ICUadmission | 0.81±0.05 | 0.80±0.09 | **0.84±0.08** | 0.79±0.10 | 0.73±0.12 |

The highest mean accuracy was obtained with two hidden layers for Algerian Death and ICU admission, and four hidden layers for Algerian Clinical status. For the French dataset, the highest mean accuracy was obtained with two hidden layers for Death and Clinical status, and three hidden layers for ICU admission. The French Death model showed equal mean accuracy with two and three hidden layers (0.89).

The standard deviations indicate variability across repeated runs, reaching 0.17 for the Algerian Clinical model with five hidden layers. In addition, the Algerian ICU admission models had mean sensitivity below 0.25 across all hidden-layer configurations. This indicates that their relatively high accuracy values did not necessarily reflect effective identification of patients in the positive class.

### 4.4 Feature rankings

Feature importance was examined for the selected ML models, while SHAP values were used to assess feature contributions in the MLP models. The results for all three prediction outcomes in both datasets are presented in Figures 8 and 9 in the Appendix, respectively. Table 7 summarises the leading feature identified for each model.

**Table 7:**
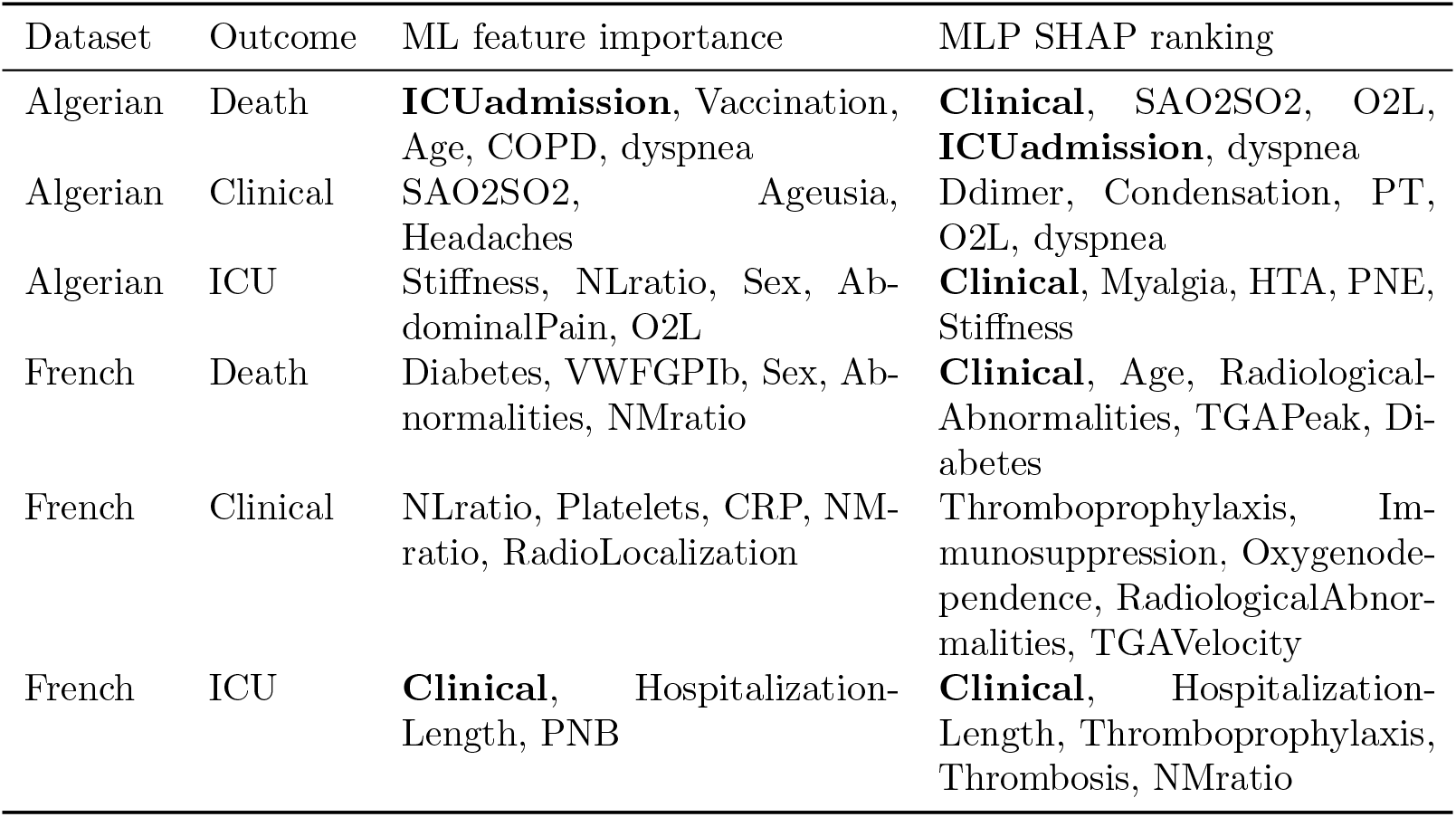
Highest-ranked features for each dataset and prediction outcome, in descending order of importance.

| Dataset | Outcome | ML feature importance | MLP SHAP ranking |
|---|---|---|---|
| Algerian | Death | **ICUadmission**, Vaccination, Age, COPD, dyspnea | **Clinical**, SAO2SO2, O2L, **ICUadmission**, dyspnea |
| Algerian | Clinical | SAO2SO2, Ageusia, Headaches | Ddimer, Condensation, PT, O2L, dyspnea |
| Algerian | ICU | Stiffness, NLratio, Sex, Ab-dominalPain, O2L | **Clinical**, Myalgia, HTA, PNE, Stiffness |
| French | Death | Diabetes, VWFGPIb, Sex, Ab-normalities, NMratio | **Clinical**, Age, Radiological-Abnormalities, TGAPeak, Di-abetes |
| French | Clinical | NLratio, Platelets, CRP, NM-ratio, RadioLocalization | Thromboprophylaxis, Im-munosuppression, Oxygenode-pendence, RadiologicalAbnor-malities, TGAVelocity |
| French | ICU | **Clinical**, Hospitalization-Length, PNB | **Clinical**, Hospitalization-Length, Thromboprophylaxis, Thrombosis, NMratio |

The most important features varied across datasets, prediction outcomes, and modelling approaches. For the ML models, the leading features were ICUadmission, SAO2SO2, and Stiffness for the Algerian Death, Clinical status, and ICU admission models, respectively. For the French models, they were Diabetes, NLratio, and Clinical, respectively.

For the MLP models, SHAP identified Clinical, Ddimer, and Clinical as the leading features for the Algerian Death, Clinical status, and ICU admission models, respectively. In the French dataset, the leading features were Clinical, Thromboprophylaxis, and Clinical, respectively.

Clinical status and ICU admission were also retained as predictors in models for other outcomes because they were considered clinically relevant to the prediction targets. Overall, feature rankings differed according to the dataset, prediction outcome, and modelling approach.

### 4.5 ML versus DL

Table 8 compares the selected ML models with the MLP configurations that achieved the highest mean accuracy over 30 runs. The ML models achieved higher test accuracy in five of the six tasks. For Algerian Clinical status, both approaches showed relatively low accuracy (0.47 for ML and 0.51 for DL).

**Table 8:** Comparison of the selected ML models and the best-performing MLP configurations across the three prediction outcomes in the Algerian and French datasets.

|  |  | ML |  |  |  | DL (MLP) |  |  |  |
|---|---|---|---|---|---|---|---|---|---|
| Dataset | Outcome | Acc. | Sens. | Spec. | F1 | Acc. | Sens. | Spec. | F1 |
| Algerian | Death | 0.92 | 1.00 | 0.90 | 0.86 | 0.81 (2) | 0.64 | 0.90 | 0.52 |
| Algerian | Clinical | 0.47 | 0.60 | 0.20 | 0.60 | 0.51 (4) | 0.62 | 0.20 | 0.62 |
| Algerian | ICU | 0.86 | 1.00 | 0.83 | 0.67 | 0.81 (2) | 0.14 | 0.93 | 0.14 |
| French | Death | 0.90 | 1.00 | 0.89 | 0.67 | 0.89 (2) | 0.40 | 0.94 | 0.40 |
| French | Clinical | 0.90 | 0.85 | 1.00 | 0.92 | 0.76 (2) | 0.84 | 0.71 | 0.81 |
| French | ICU | 0.95 | 0.83 | 1.00 | 0.91 | 0.84 (3) | 0.82 | 0.92 | 0.80 |

Differences were also observed in sensitivity, particularly for Algerian ICU admission (1.00 for ML versus 0.14 for DL) and French Death (1.00 versus 0.40). However, this comparison should be interpreted cautiously because ML performance was measured on a single test set, whereas DL results represent averages across 30 runs.

### 4.6 Friedman test

The Friedman test was used to compare the 15 classifier–feature-selection combinations within each dataset (Fig. 6). In both the Algerian and French datasets, CART combined with ANOVA achieved the highest rank, followed by LR with RFE and LR with ANOVA. The lowest-ranked combinations were NB with MI and NB with RFE. These results indicate that CART with ANOVA ranked first across both datasets.

**Figure 6:**
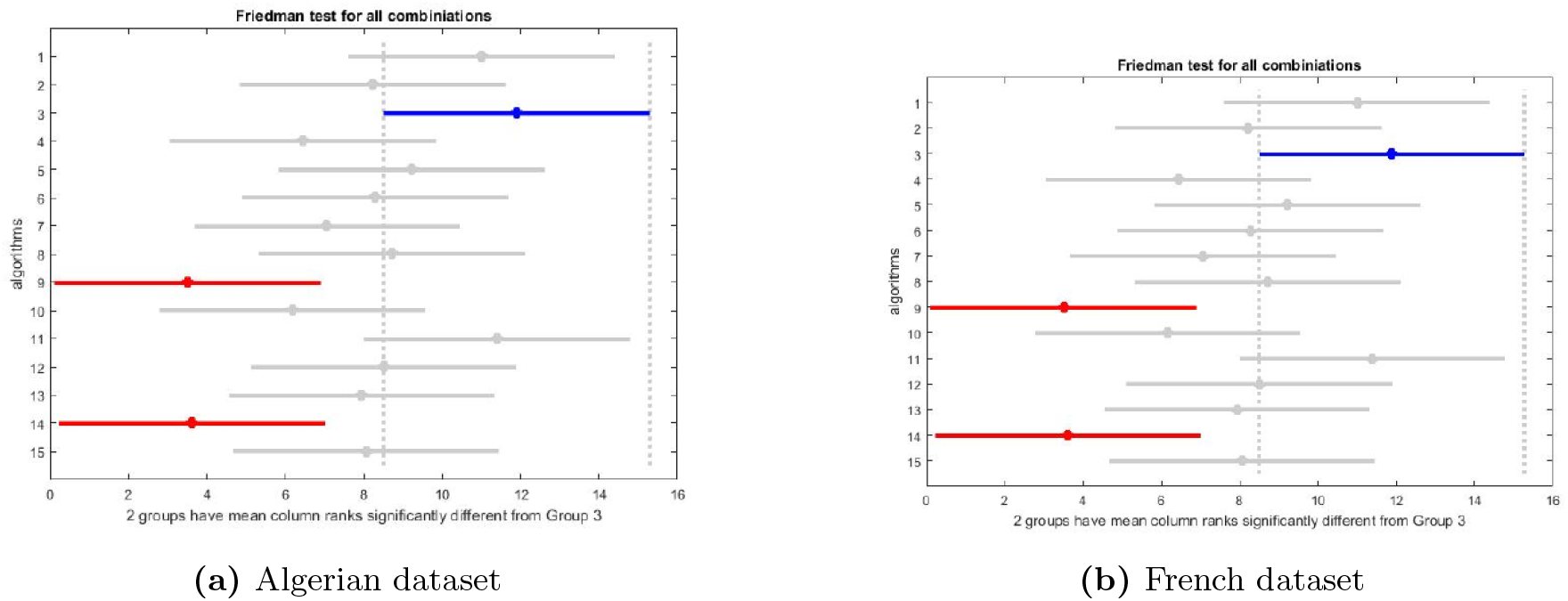
Friedman test rankings of the 15 classifier–feature-selection combinations for the Algerian and French datasets.

## 5 Discussion

### 5.1 Interpretation of findings

The results showed that model performance depended on the dataset and prediction outcome. Although selecting 20 features generally improved cross-validation performance, no single classifier–feature-selection combination was best for all six tasks. The Friedman analysis identified CART with ANOVA as the highest-ranked combination in both datasets.

The selected ML models achieved higher hold-out accuracy than the average MLP models in five of the six tasks. However, this comparison should be interpreted cautiously because ML performance was evaluated on one test set, whereas DL performance was averaged over 30 runs.

Performance also varied by outcome. Death and ICU admission generally showed higher accuracy, but the test sets contained few positive cases. The Algerian ICU MLP models, in particular, had low sensitivity. Clinical-status prediction was more challenging in the Algerian dataset than in the French dataset.

### 5.2 Scientific implications

This study highlights the importance of evaluating different combinations of classifiers and feature-selection methods for each prediction outcome and dataset. It also provides a comparative framework that can be further evaluated using larger patient cohorts. In addition, the Algerian dataset contributes data from a setting that is less represented in COVID-19 prognostic research.

### 5.3 Strengths and limitations

#### 5.3.1 Strengths

The main strengths of this study include:

- The use of two datasets from different countries (Algeria and France).
- The investigation of three clinically relevant outcomes: death, ICU admission, and clinical status.
- The comparison of five machine learning classifiers and 15 classifier–feature-selection combinations.
- The evaluation of multilayer perceptron (MLP) models with different numbers of hidden layers over 30 runs to account for variability between runs.
- The consideration of clinical input during model selection and interpretation.

#### 5.3.2 Limitations

Despite these strengths, several limitations should be acknowledged:

- **Small sample sizes:** The datasets included 80 and 99 patients, respectively, and the test sets contained only 13–20 patients. Therefore, the classification of a single patient could substantially affect the reported performance metrics.
- **Model selection:** MLP configurations were selected based on test-set accuracy, which may have led to optimistic performance estimates.
- **Class imbalance:** The death and ICU admission outcomes were imbalanced, and no specific method was applied to address this issue.
- **External validation:** The models were not evaluated on independent external datasets, which limits the generalizability of the findings.

Overall, these limitations indicate that the results should be considered exploratory. Further evaluation using larger, independent datasets is needed before considering potential clinical applications.

## 6 Conclusion and Future Directions

### Conclusion

This study investigated the use of machine learning (ML) and deep learning (DL) methods to predict three outcomes in hospitalised COVID-19 patients: death, ICU admission, and clinical status. The experiments were conducted using two datasets from Algeria and France.

The main findings can be summarised as follows:

- Five ML classifiers were evaluated alongside three feature-selection methods and multilayer perceptron (MLP) models with different numbers of hidden layers.
- Selecting 20 features generally improved cross-validated accuracy compared with using 30 or all features.
- The best-performing ML model varied according to the dataset and prediction outcome.
- CART combined with ANOVA achieved the highest rank in the Friedman test for both datasets.
- The selected ML models achieved higher hold-out accuracy than the average MLP models in five of the six prediction tasks.
- The important features varied across datasets, prediction outcomes, and modelling approaches.

However, the small sample sizes and methodological limitations mean that these findings should be considered exploratory. The models are not yet ready for clinical use and require further validation.

### Future Work

The following directions are proposed to improve and extend this study.

**Improvements Proposed in the Thesis**

- **Expand the datasets:** Add more medical records to the Algerian dataset to increase the number of available observations.
- **Improve the MLP architecture:** Experiment with the number of neurons in each layer, the number of training epochs, and dataset size.
- **Tune ML models:** Adjust classifier hyperparameters to improve model performance.
- **Evaluate additional methods:** Introduce other classification algorithms and feature-selection methods and compare their performance.

**Additional Methodological Recommendations**

- Apply nested cross-validation and perform preprocessing within each training fold.
- Reassess predictors that may be derived from or closely related to the prediction outcomes.
- Report confidence intervals and evaluate probability calibration.
- Perform external validation using independent datasets.
- Investigate data-augmentation methods to address limited sample sizes and class imbalance.

## Declarations

### Origin of the work

This preprint is based on the Master’s thesis “Application of Machine Learning and Deep Learning in diagnosis Covid19” (L. Merchi and C. Boucheham; supervisor A. Layeb; co-supervisor A. Derouiche), University of Abdelhamid Mehri – Constantine 2, June 2022.

### Ethics and data governance

The Algerian data were extracted from the archive of the internal medicine department of UCH Constantine with the permission of the head of department.

### Data and code availability

The Algerian dataset and the analysis code are not publicly available. The French dataset was obtained from the published study by Billoir et al. (2021).

## Acknowledgements

The authors thank Prof. Y. Kitouni, head of the internal medicine department at UCH Constantine, for access to the archive, and Dr. A. Derouiche for guidance.

## A Dataset attribute listings

**Table 9:**
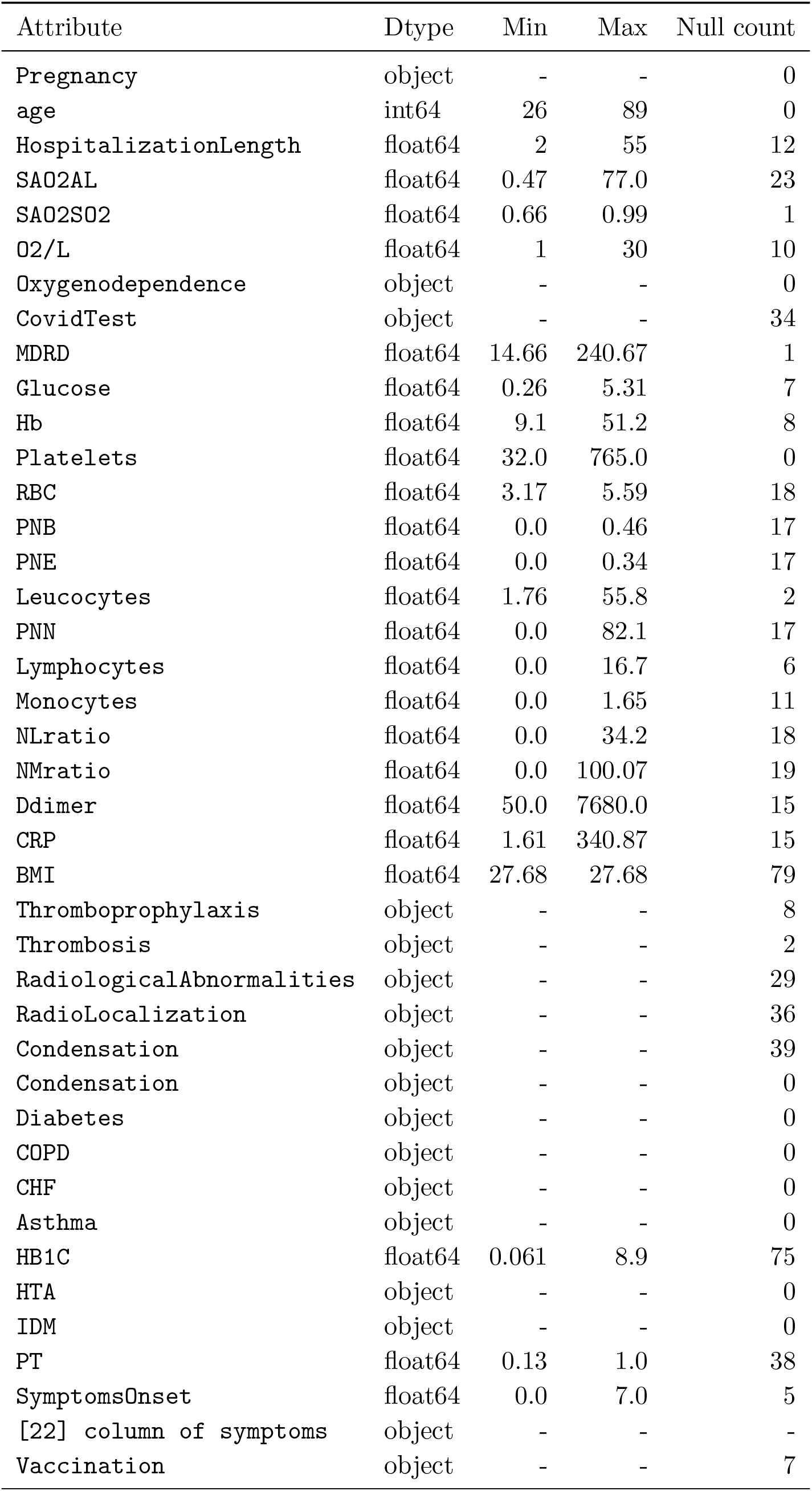
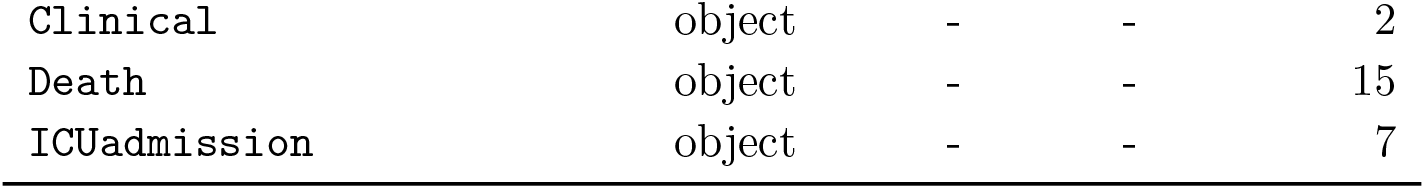
Attributes of the Algerian dataset.

| Attribute | Dtype | Min | Max | Null count |
|---|---|---|---|---|
| Pregnancy | object | - | - | 0 |
| age | int64 | 26 | 89 | 0 |
| HospitalizationLength | float64 | 2 | 55 | 12 |
| SAO2AL | float64 | 0.47 | 77.0 | 23 |
| SAO2SO2 | float64 | 0.66 | 0.99 | 1 |
| O2/L | float64 | 1 | 30 | 10 |
| Oxygenodependence | object | - | - | 0 |
| CovidTest | object | - | - | 34 |
| MDRD | float64 | 14.66 | 240.67 | 1 |
| Glucose | float64 | 0.26 | 5.31 | 7 |
| Hb | float64 | 9.1 | 51.2 | 8 |
| Platelets | float64 | 32.0 | 765.0 | 0 |
| RBC | float64 | 3.17 | 5.59 | 18 |
| PNB | float64 | 0.0 | 0.46 | 17 |
| PNE | float64 | 0.0 | 0.34 | 17 |
| Leucocytes | float64 | 1.76 | 55.8 | 2 |
| PNN | float64 | 0.0 | 82.1 | 17 |
| Lymphocytes | float64 | 0.0 | 16.7 | 6 |
| Monocytes | float64 | 0.0 | 1.65 | 11 |
| NLratio | float64 | 0.0 | 34.2 | 18 |
| NMratio | float64 | 0.0 | 100.07 | 19 |
| Ddimer | float64 | 50.0 | 7680.0 | 15 |
| CRP | float64 | 1.61 | 340.87 | 15 |
| BMI | float64 | 27.68 | 27.68 | 79 |
| Thromboprophylaxis | object | - | - | 8 |
| Thrombosis | object | - | - | 2 |
| RadiologicalAbnormalities | object | - | - | 29 |
| RadioLocalization | object | - | - | 36 |
| Condensation | object | - | - | 39 |
| Condensation | object | - | - | 0 |
| Diabetes | object | - | - | 0 |
| COPD | object | - | - | 0 |
| CHF | object | - | - | 0 |
| Asthma | object | - | - | 0 |
| HB1C | float64 | 0.061 | 8.9 | 75 |
| HTA | object | - | - | 0 |
| IDM | object | - | - | 0 |
| PT | float64 | 0.13 | 1.0 | 38 |
| SymptomsOnset | float64 | 0.0 | 7.0 | 5 |
| [22] column of symptoms | object | - | - | - |
| Vaccination | object | - | - | 7 |
| Clinical | object | - | - | 2 |
| Death | object | - | - | 15 |
| ICUadmission | object | - | - | 7 |

**Table 10:** Attributes of the French dataset.

| Attribute | Dtype | Min | Max | Null count |
|---|---|---|---|---|
| Age | int64 | 19.0 | 94.0 | 1 |
| Sex | object | - | - | 1 |
| SymptomsOnset | float64 | 1.0 | 23.0 | 3 |
| PCRCoVID | object | - | - | 1 |
| Oxygenodependence | object | - | - | 1 |
| Tobacco | object | - | - | 1 |
| RadiologicalAbnormalities | object | - | - | 14 |
| RadioLocalization | object | - | - | 19 |
| Condensation | object | - | - | 18 |
| Immunosuppression | object | - | - | 0 |
| BMI | float64 | 17.0 | 40.1 | 9 |
| HTA | object | - | - | 1 |
| Diabetes | object | - | - | 1 |
| Respiratory | object | - | - | 1 |
| COPD | object | - | - | 1 |
| Asthma | object | - | - | 1 |
| Other | object | - | - | 1 |
| MDRD | float64 | 16.41597543 | 280.8168476 | 1 |
| CHF | object | - | - | 1 |
| HospitalizationLength | float64 | 1.0 | 58.0 | 2 |
| Thromboprophylaxis | object | - | - | 1 |
| Thrombosis | object | - | - | 1 |
| RBC | float64 | 2.66 | 6.78 | 2 |
| Hb | float64 | 7.0 | 17.3 | 2 |
| Platelets | float64 | 56.0 | 718.0 | 2 |
| Leukocytes | float64 | 1.5 | 109.0 | 2 |
| PNN | float64 | 0.68 | 26.71 | 2 |
| PNE | float64 | 0.0 | 1.89 | 2 |
| PNB | float64 | 0.0 | 0.25 | 2 |
| Lymphocytes | float64 | 0.3 | 34.2 | 2 |
| Monocytes | float64 | 0.13 | 1.42 | 2 |
| NLratio | float64 | 0.35 | 43.79 | 2 |
| NMratio | float64 | 0.2 | 56.12 | 2 |
| PT | float64 | 11.6 | 46.7 | 1 |
| aPTT | float64 | 0.83 | 1.95 | 3 |
| Ddimer | float64 | 174.0 | 20000.0 | 1 |
| Antithrombin | float64 | 41.0 | 132.0 | 1 |
| FibrinMonomer | float64 | 0.67 | 400.0 | 1 |
| Fibrinogen | float64 | 2.17 | 11.98 | 1 |
| VWFGPIb | float64 | 35.0 | 722.0 | 1 |
| ProthrombinFragment1+2 | float64 | 41.0 | 2177.0 | 1 |
| DICDdimer | float64 | 0.0 | 5.0 | 1 |
| DICFibrinMonomer | float64 | 0.0 | 5.0 | 1 |
| TGAlagtime | float64 | 2.92 | 17.5 | 3 |
| TGAETP | float64 | 282.0 | 2046.29 | 3 |
| TGAPeak | float64 | 31.14 | 380.12 | 3 |
| TGATTP | float64 | 5.42 | 25.83 | 3 |
| TGAVelocity | float64 | 3.97 | 145.69 | 3 |
| CRP | float64 | 5.0 | 343.0 | 20 |
| ICUadmission | object | - | - | 1 |
| Clinical | object | - | - | 0 |
| Death | object | - | - | 0 |

## B Additional figures

### ROC curves, selected ML models

**Figure 7:**
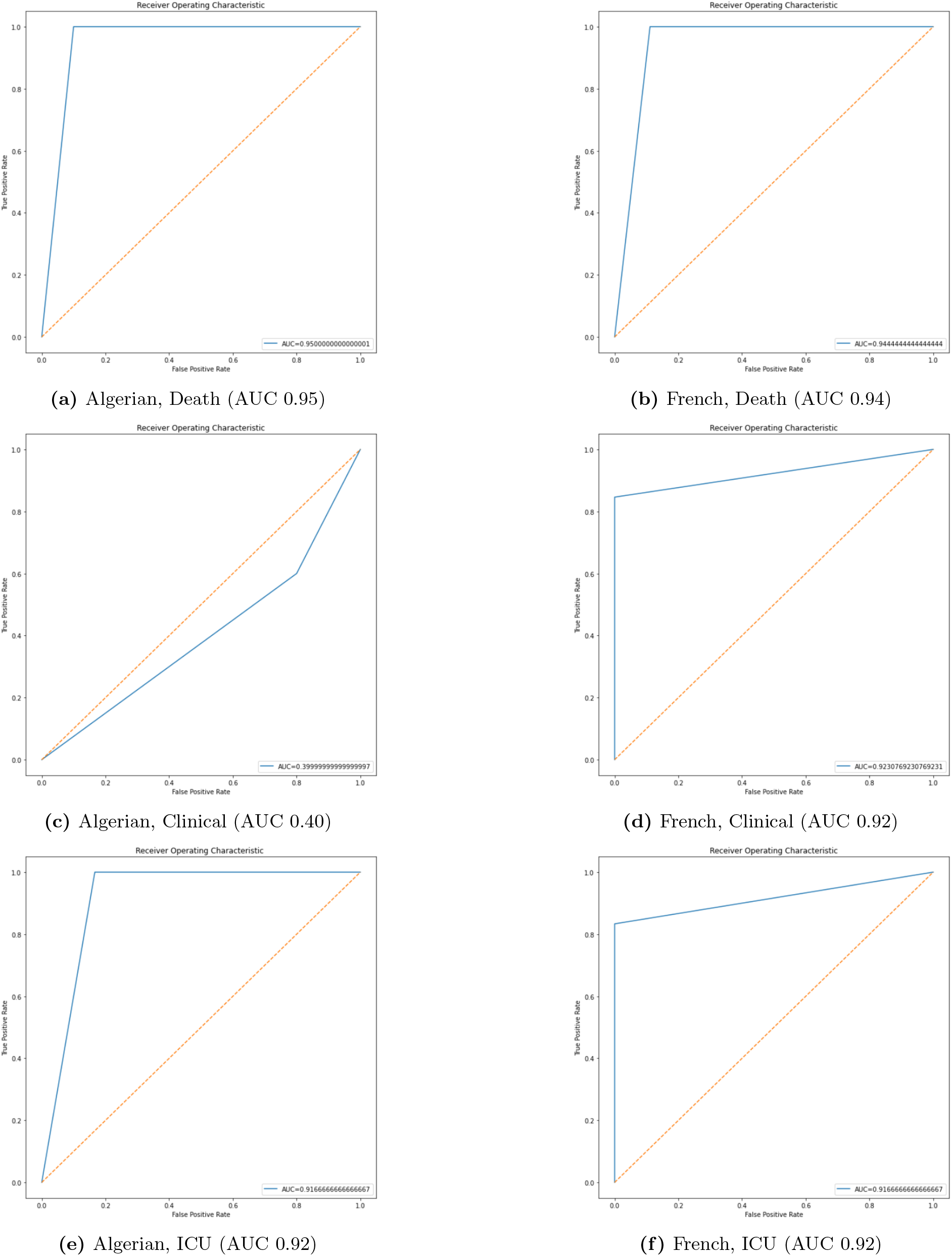
ROC curves of the selected ML models on the test sets.

### Feature importance, selected ML models

**Figure 8:**
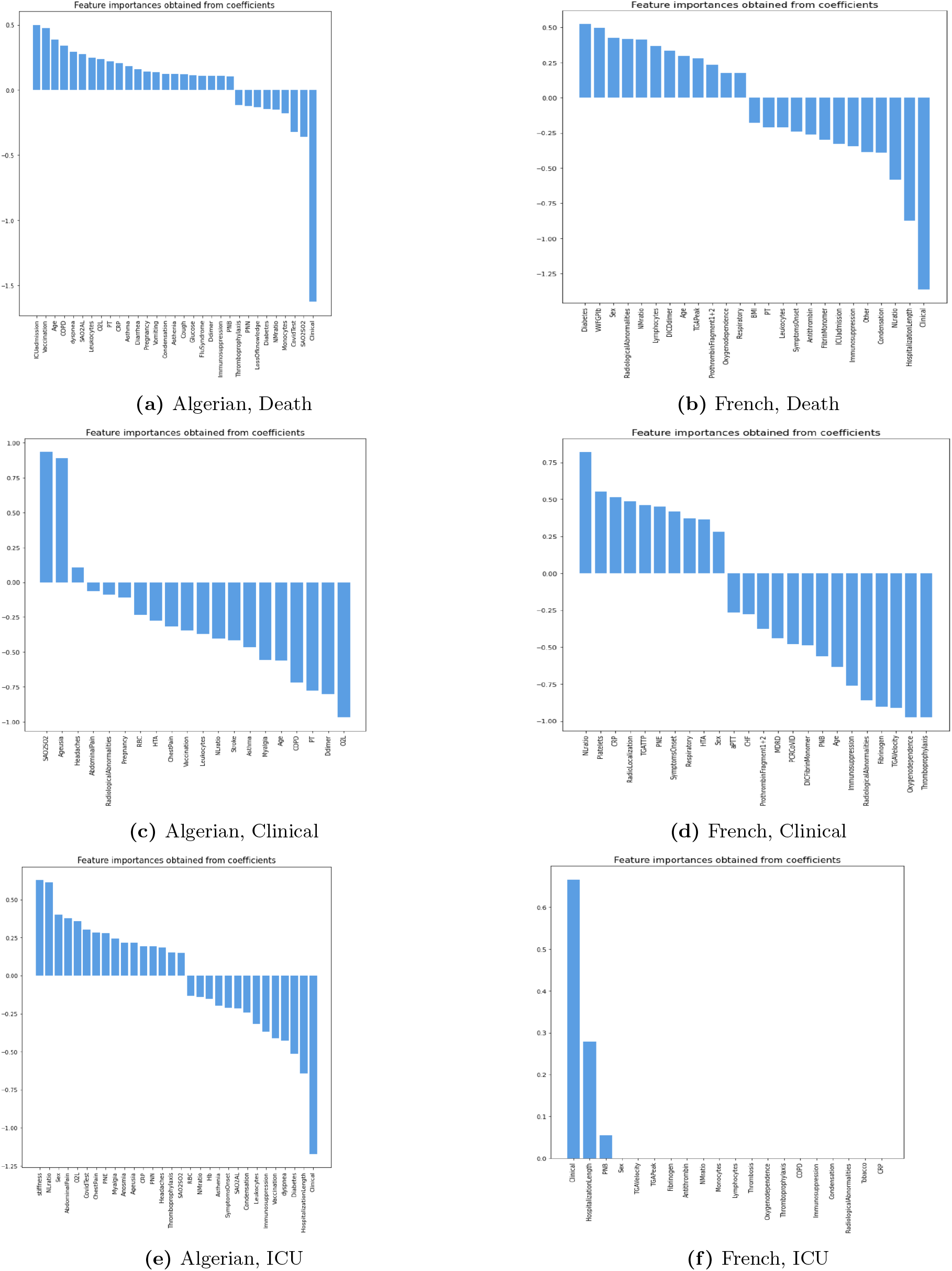
Feature-importance scores of the selected ML models.

### SHAP summaries, MLP models

**Figure 9:**
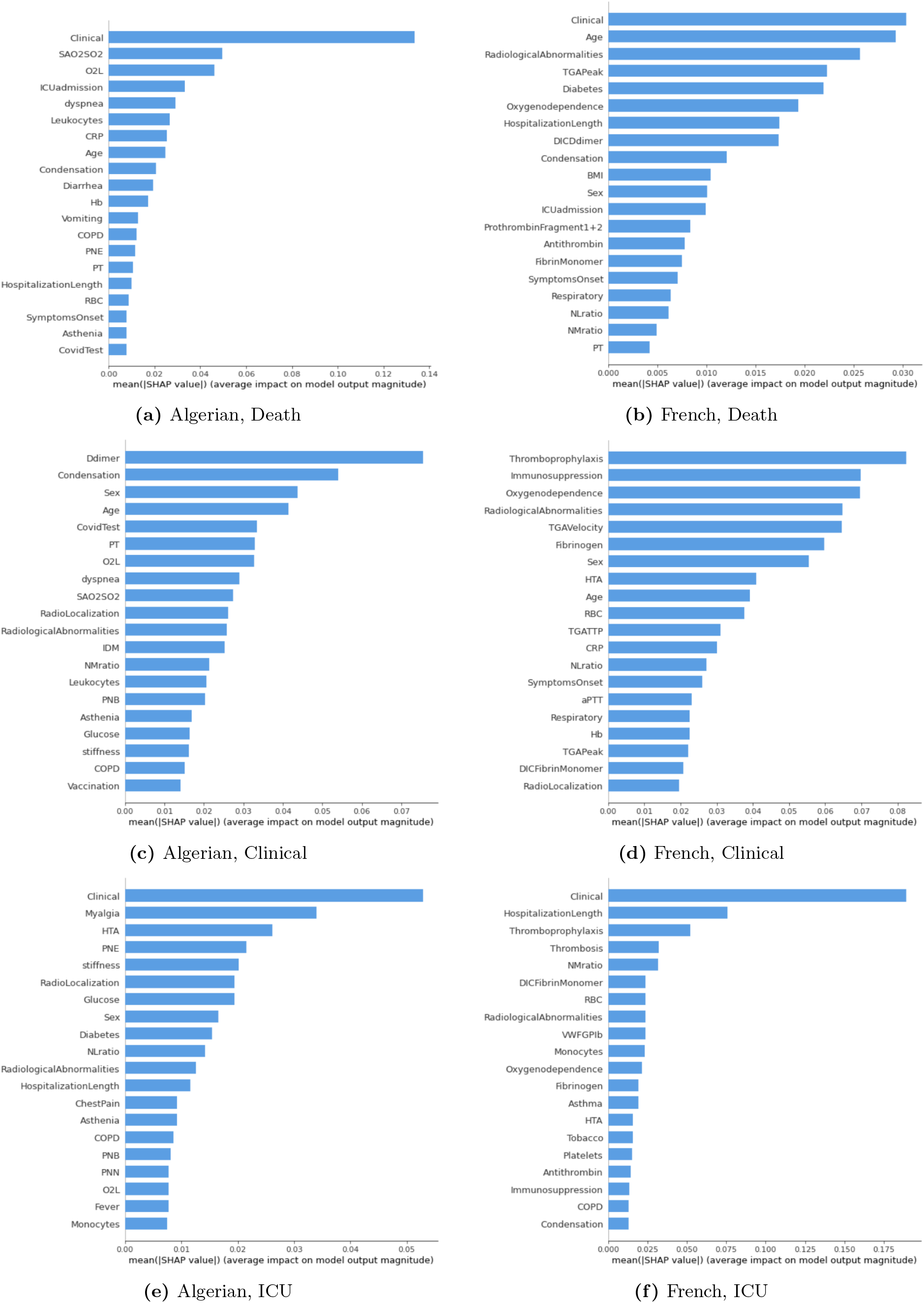
Mean absolute SHAP values of the best MLP of each task.

### MLP architectures

**Figure 10:**
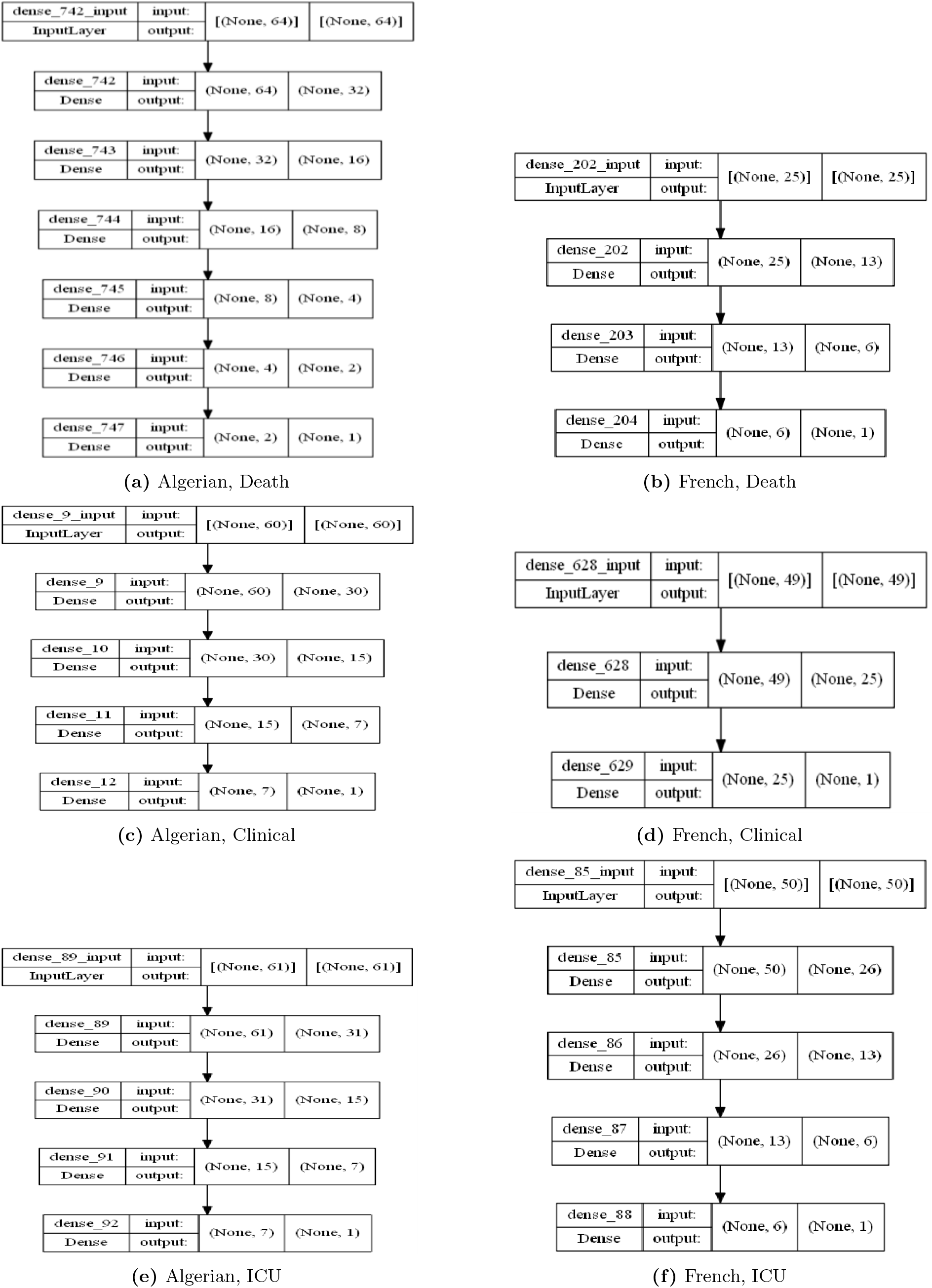
Architectures of the best single-run MLP for each task.

## Notes

### Competing Interest Statement

The authors have declared no competing interest.

### Author Declarations

The Algerian dataset was developed in collaboration with the Internal Medicine Department of the University Hospital Centre of Constantine (UCH Constantine), Algeria. Patient data were extracted from the department's medical archives under the supervision of Dr. Asma Derouiche, following authorization from Prof. Y. Kitouni, Head of the Internal Medicine Department, who granted access to the confidential patient records. This dataset was developed as part of the authors' Master's thesis research.

